# Healthcare Facility Water, Sanitation and Hygiene Service Status and Barriers in Ethiopia: It’s Implication for COVID-19 pandemic and Healthcare Acquired Infection Prevention

**DOI:** 10.1101/2022.12.09.22283296

**Authors:** Atimen Derso, Taffere Addis, Bezatu Mengistie, Awoke keleb, Ayechew Ademas

**Affiliations:** Institute of Ethiopian Water Resource, Addis Ababa University, Addis Ababa, Ethiopia; Department of Environmental Health, College of Medicine and Health Sciences, Wollo University, Dessie, Ethiopia

**Keywords:** WASH services, Healthcare facilities, Barriers, COVID-19, Ethiopia

## Abstract

**Background:** Despite the public health significance of healthcare Water, Sanitation, and Hygiene (WASH) service in reduction of nosocomial infection and improving quality of care is paramount little is known on the status of WASH service in a health care facility at the time of pandemic and the barriers that hinder the service in the health care setting in Ethiopia.

**Objective:** The aim of this study was to assess status of basic water, sanitation, hand hygiene, healthcare waste management, and environmental cleanliness service and its barriers at public health care facilities in the city of Addis Ababa, Ethiopia 2022.

**Methods:** Convergent parallel mixed design was conducted among 86 public health care facilities located in Addis Ababa city. Stratified sampling technique was used to select health care facilities. A semi-structured observational checklist tool was used to measure the availability of services. For the qualitative study, semi-structured interview was conducted among 16 key informants and thematic data analysis was done to identify the barriers.

**Finding:** This study found that no one healthcare facility had basic access to overall WASH services. The independent WASH domain analysis showed that, about 86% healthcare facilities had basic water access, 100% had limited sanitation access, 88.4% had limited hand hygiene service, 69.8% had limited healthcare waste management service, and 97.7% had limited environmental cleaning service. Built environments of WASH infrastructure; Resource availability and allocation; leadership and stakeholder participation; inadequate training and poor behaviour; and legal issues were identified barriers to provision of basic healthcare WASH services.

**Conclusion and recommendation:** The availability of healthcare WASH services in Addis Ababa city remains far from the pace to achieve the sustainable goal target by 2025. The limited access to WASH services makes worsening the prevention and control of COVID-19 pandemics, healthcare acquired infection in the facility. The country need to act now on more financial investment, capacity building, facilitating committed leadership, and participation of stakeholders to ensuring basic WASH services at healthcare setting.

## Introduction

Availability of sustainable WASH in a health care facility is critical for quality care to meet individual preferences, needs, and values; without minimum standards of WASH service in all health care facilities cannot meet the demand for good quality of care and human right to health (1). Provision of basic WASH service is a prerequisite for infection prevention and control(IPC) which can protect front-line health care workers, patients, visitors from infectious disease transmission in a health care setting; and it is human rights, dignity, social justice and gender issue at any health care setting (1, 2).

Inadequate WASH remains one of the determinants of the global burden of disease as pointed out by Pruss-Ustun et.al (3). Lack of basic healthcare WASH services is a major public health problem, and it continues a challenge to international communities. Emerging and re-emerging diseases like Antimicrobials Resistance (AMR), Ebola, and COVID-19 pandemic place WASH as a centre of disease prevention. Health care WASH is more important than ever in the role of prevention of COVID-19 and AMR among patients, staff, or community (4, 5). However, worldwide 11% of healthcare facilities had no water service, 10% of health care facilities had no sanitation service, and only 51% of health care facilities had basic hand hygiene service at the point of care and nearby toilet by 2022, which makes hundreds of millions of people are face risk of infection; since more than 681 million people lacked basic waste management service in health care facilities in Sub-Sahara African countries, and 3.85 billion people worldwide lacked basic hand hygiene service at their healthcare facilities (6).

The Lancet review reveals that healthcare-associated infection burden is much higher in developing than developed countries, which is more than 15% of patients developed an infection when they are staying in hospitals; and the risk of patients admitted to health care facility acquire one or more infection is two to twenty times higher developing than developed countries (7, 8). Systematic review in Africa reveal that poor WASH services provision in the HCF causes women to choose home delivery and increase patient dissatisfaction in low and middle-income countries (9). We argue that health care professionals are at the front line of protecting clients from infection and covid-19 pandemic response, and they are exposed to infection when they are working in a high-risk environment. As reported by Desta et.al (10), the availability of supplies and infrastructure of WASH service is critical to safeguard health care professionals and to enhance the practice of IPC. The lesson gain from Ebola outbreak and Severe Acute Respiratory Syndrome (SARS) implies poor WASH service in the health system ready-made to expose clients and frontline workers to infection and expand the COVID-19 pandemic. Worldwide the proportion of health care workers infected with SARS was from 20% to 60% (11); and Cooper et.al reported, there was challenging to expand the outbreak of Ebola in Liberian healthcare facilities with poor WASH services for IPC (12).

To improve WASH service in a health care facility, which has public health significance in the reduction of nosocomial infection, improving quality of care, increasing healthcare-seeking behaviour, and averting cost expend for infection Ethiopia launched WASH initiatives, clean and safe health facility (CASH), and sector-wide One-WASH national program (OWNP) (13). However, there is a dearth of evidence on the status of basic health care WASH service and the barriers that setback the provision of WASH services in a health care setting in Ethiopia. Therefore the aim of this study was to assess the status of basic WASH services availability and explore the barriers to providing adequate WASH services in public health care setting through mixed research design in the city of Addis Ababa, Ethiopia.

## Materials and Methods

### Study Area

The study was conduct in Addis Ababa city. There are 14 public hospitals including specialty centres and COVID-19 treatment centres, and 100 health centres located in the city of Addis Ababa. With the efforts of private health care facility these public health care facilities are serving in a range of health care services for Addis Ababa city population and part of Ethiopia, which is estimated to be around 5,228,000 by 2022 (https://population.un.org/wpp/). During pandemic, Addis Ababa city is the city where the highest number of COVID-19 cases located which accounting 54.2% (254,447) of people tested positive were located in Addis Ababa city from the total of 469,581 positive cases. (https://ethiopianhealthdata.org/dashboard/covid19-ethiopia). (Figure 1)

**Figure 1:**
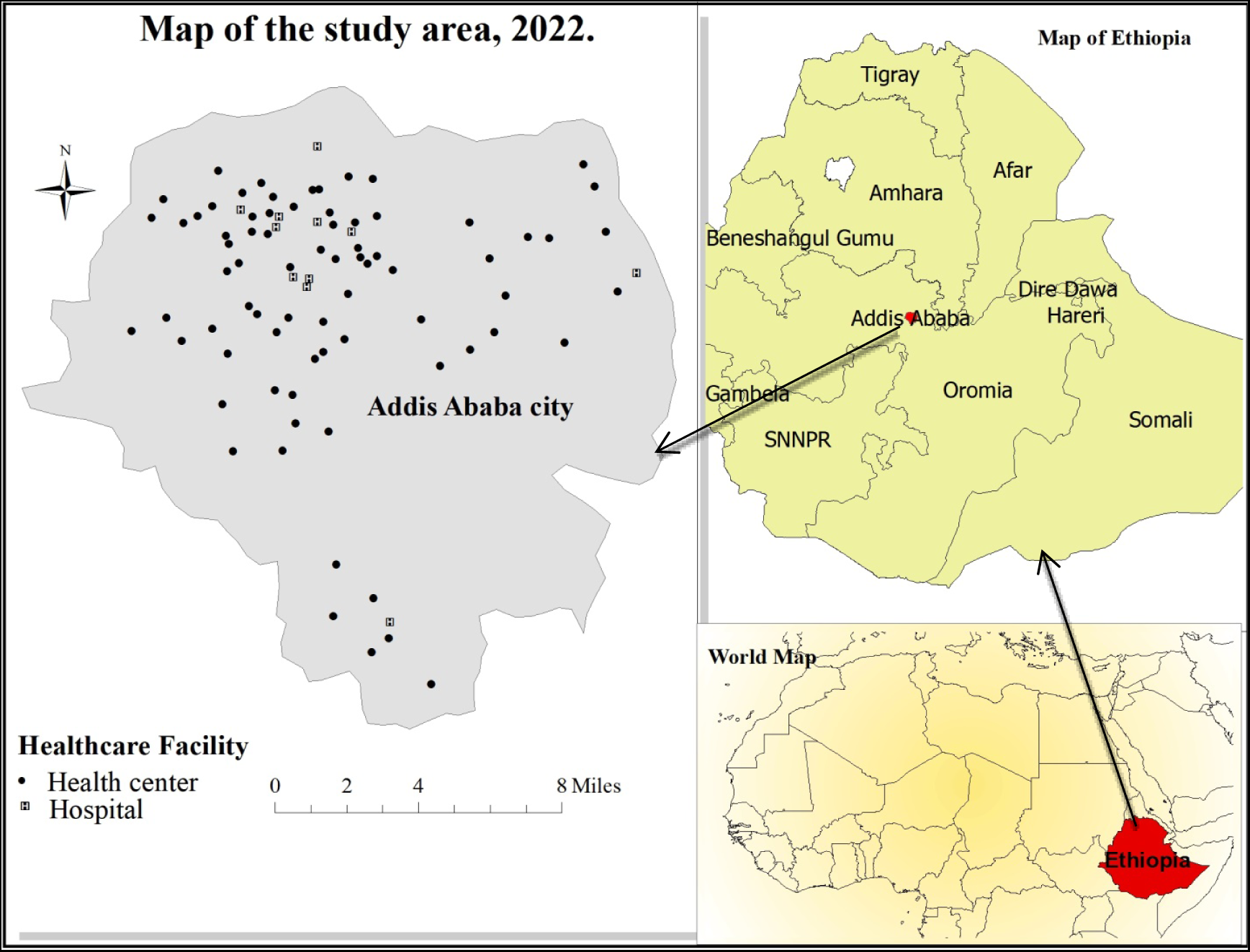
Spatial location of sampled healthcare facility

### Study Design/Approach and period

Convergent parallel mixed design was conducted. We intend to use convergent parallel mixed design to collect both quantitative and qualitative forms of data independently at the same time farm on the same concepts of WASH services and analyse and integrate the information to final interpretation of results to have a complete understanding of the research questions(14). The study was conducted from March, 2022 to June, 2022.

### Study population

The study population of this study was all public health care facilities, health centre, specialty centres, and referral and teaching hospitals located in the city of Addis Ababa.

### Inclusion and Exclusion criteria

**Inclusion criteria**: public healthcare facilities in Addis Ababa which provide service for the communities during the study were included.

**Exclusion criteria:** public healthcare facility in the city which is exclusively serving for a COVID-19 treatment centre was excluded.

### Study Variables

Dependent variable: WASH services status

Independent variables: WASH service barriers

## Operational Definition

**WASH services status:** The status of WASH services in the health care facility will be measured by JMP health care facility WASH standard, and the five indicators of WASH service has been classified into separate three-level service ladder (basic, limited, and no service) as guided by GMP standard (16).

**Basic Water Service:** health care facility where the main source of water is an improved source and located on the premises, and from which water is available at the time of the survey (16).

**Basic Sanitation Service:** health care facility with improved and usable sanitation facilities, and with at least one toilet dedicated for staff, and one sex separated toilet with menstrual hygiene facilities and one toilet accessible for users with limited mobility (16).

**Basic Hand Hygiene Service:** health care facility with fictional hand hygiene facilities are available at one or more points of care and within 5 meters of toiles (16).

**Basic Health Care Waste Management Services:** healthcare facilities where waste is safely segregated in consultation areas, and sharps wastes are treated and disposed of safely, and infectious wastes are treated and disposed of safely (16).

**Basic environmental cleaning services:** health care facility that have a protocol for cleaning, and staff with cleaning responsibilities have all received training on cleaning procedures (16).

**Barriers:** barriers are challenges or bottlenecks that deter health care facilities to provide adequate WASH service or to the improvement of WASH services in health care facility (17).

### Sample Size Determination and Sampling Procedure for quantitative study

Sample size was determined using a single proportion formula with the assumption of the estimated availability of basic treatment and disposal of healthcare waste service in Ethiopia was 64 % from JMP baseline healthcare WASH report (18), 95% confidence interval (CI) and α=5%, 5% marginal error.

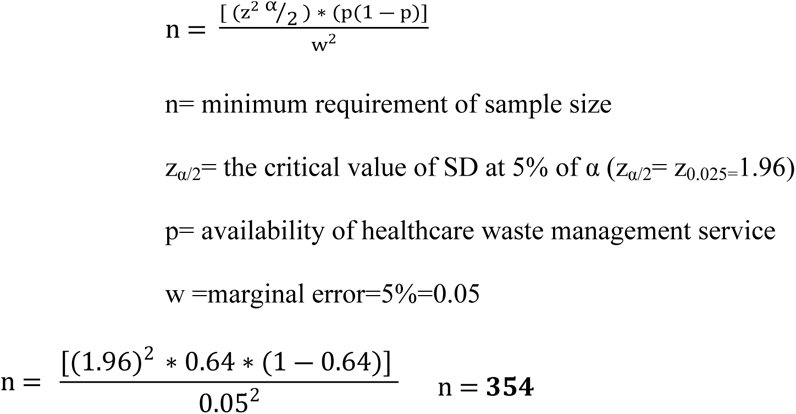

However, the total source population in this study is 114 healthcare facility which less than 10,000. Based on this population we used reduction formula to calculate final sample size as recommended by scholars.

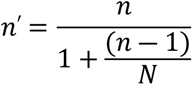

*n*′= final sample size, *n* = first calculated sample size, *N* = total study/source population

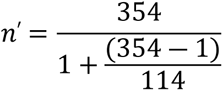

*n*′= <u>86,</u> the study need 86 healthcare facilities.

To get representative sample stratified random sampling technic was used. Simple random sampling technique was applied to select hospital and health centre.

### Study participants and recruitment technic for Qualitative study

For the qualitative study, there are no agreed ways to determine the sample size (15). The saturation or redundancy of information about all need concepts after conducting a sequential interview was determine the number of sample size (19). Accordingly, the study participant, 16 key informants, were selected through purposive sampling method from hospitals (11 participants IPC focal person) and health center (4 medical directors), and Addis Ababa health bureau (one WASH program expert) based on personal experience or knowledge on healthcare WASH services and exposure to WASH services barriers at a health care facility and whose views or opinions can provide focused, useful, and creditable rich information..

### Data collection tool and procedure for quantitative study

The quantitative data was collected by a semi-structured observational checklist adapted from WHO and UNICEF joint monitoring program core question, and indicators in monitoring WASH service level in health care facilities (20). Observation of WASH services was carried out through observing water, sanitation, hand hygiene, and waste management facility and functioning of service at the time of observation, and asking and reviewing supportive documents and standards IPC department. The observation was held by trained two data collectors. After getting legitimacy the data collectors were taken the spatial location of facilities (geographical positioning coordinate), and visit the WASH infrastructure of the facility’s randomly selected outpatient service area and finally they were visit the hospital or health centre environment related to health care waste treatment and disposal system; sanitation service; hand washing service; and observing and asking environmental cleaning services and protocols.

### Data collection tool and procedure for the qualitative study

For qualitative data collection, face-to-face in-depth interviews with purposively selected key informants were conducted by the principal investigator through semi-structured interview guide which is adapted from relevant literatures. After getting participant consent in written and oral form, the principal investigator was conduct face-to-face dialogue with key informants. The interview was recorded by an audiotape recorder with additional key informant emphasis were held on memos by the interviewer. The interview was continued until no new information or concepts emerged for all questions (total 16 interviews has been conducted).

### Data quality assurance

The quality of the data and research process were ensured through adequate training (one-day training) for data collectors (BSc) and 2 supervisors (postgraduate) on the purpose of the study, part of questionnaire to assess WASH services, way of quality assurance, research ethics, and observing and data recording approach. The data collection was started within a week after the training. Data collectors were also supervised every day at the time of the survey.

To ensure the trustfulness and reliability of qualitative findings, the interview technique was revised in between interview periods if there is any new insight from the prior interview, key informants were encourage expressing their ideas and opinion freely at begging of interview and explain their experience on healthcare WASH service. We were check carefully the meaning (description) of each code with the data given that code. A detailed description of major themes has been carried out. Qualitative researchers were invited to review the finding and raise questions on them to improve the credibility of the result.

### Data management and analysis for quantitative study

The completeness of data was checked manually and after editing and clearing, data entry was done by using Epi data version 3.1. Then, the data was exported to SPSS version 25 for analysis. Descriptive analysis was carried out to quantitative data. All observed core questions under each WASH service indicator have been presented through frequency distribution tables, bar graphs, and narration. The percentage of water, sanitation, hygiene, waste management, and environmental cleanliness service availability across the health care facility was compute from the summation of core questions that are used to measure basic, limited, and no service level.

### Data management and analysis for the qualitative study

Thematic analysis method was conducted for the qualitative data. The analysis of qualitative data was undergoing after each interview has been ended. First, after conducting the interview, audio recorded data and filed note (memo) data were under go transcribed and translated to English and the text data was stored and located in qualitative data management Atlas ti software. Second, organize the data; Third-generating initial codes; Fourth-organized codes; after revising the identified codes, potential themes or contents were created by clustering or categorizing related codes based on their concepts and relationship. Fifth-organized main themes; themes were revised, interrelating, and categorized which includes the barriers of WASH provision in the health care setting, and finally main themes were defined and explained with quotes of key informants’ opinions on the barriers as a major finding as part of report.

### Ethical Consideration

Ethical clearance approval was sought from the ethical review board of St. Paul’s Hospital Millennium Medical College and Addis Ababa Public Health Research and Emergency Management Directorate. Verbal and written consent was obtained for each study participant before the interview. This study was address the code of research ethics on disclosing all the necessary information regarding autonomous rights, confidentiality, risk and benefit, free withdrawal at the time of interview.

## Results

### Water service availability

A total of 86 health care facilities were studied during the survey. Of which, 75(87.2) were health centers, and 11(12.8%) of facility were hospitals including specialty centers, referral, and specialized teaching hospitals. The mean daily client flow rate at hospital and health centre was 590.6 (± 541.5) and 220.6 (± 88.8), respectively. The overall proportion of basic water service availability in healthcare facilities in Addis Ababa was 74 (86%), while limited water service was 12(14%). At the time of the survey, one out of six health centers (16%) in Addis Ababa city had limited water service (Figure 2).

**Figure 2:**
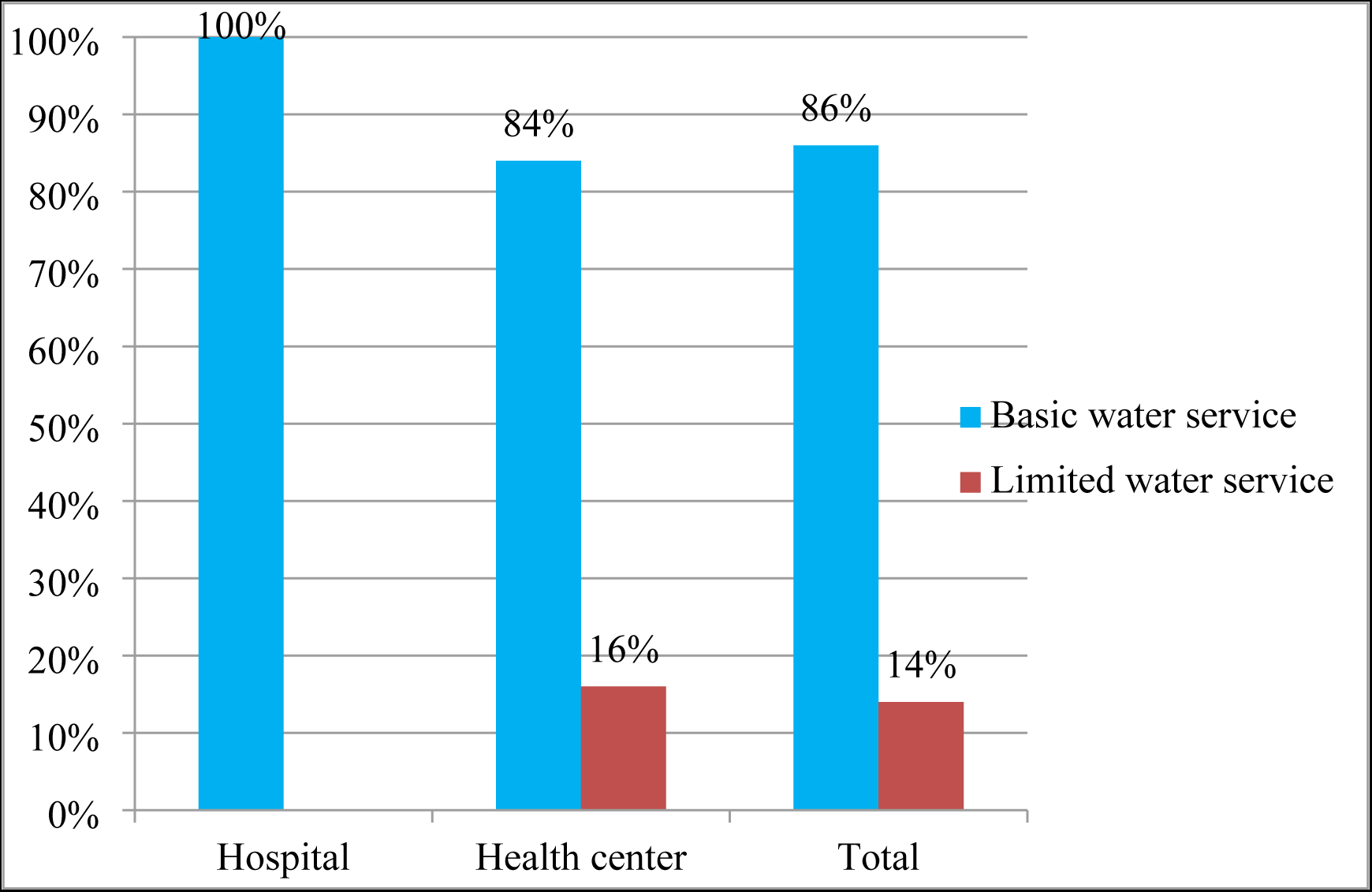
Water service availability among public healthcare facility in Addis Ababa city, Ethiopia 2022.

All studied healthcare facilities had access piped water from an improved source located within the health care facility premises. However, water was not available from main source at time of the survey at 12(14%) healthcare facility and nearly three-quarters 64(74.4%) of healthcare facilities had faced water discontinuity before the survey. Moreover, 47(54.7 %) of healthcare facility had not piped water access at the OPD (Table 1).

**Table 1:**
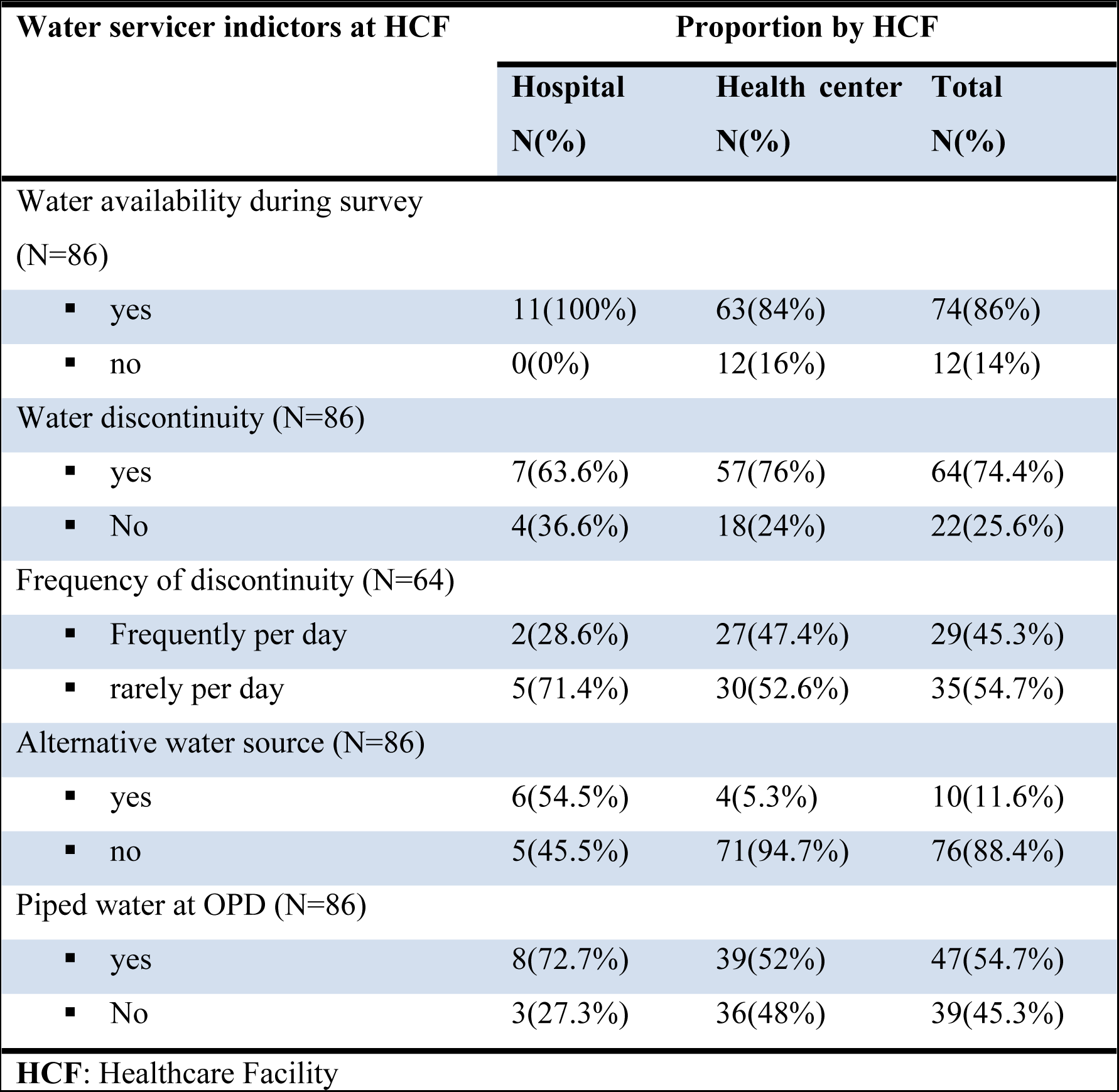
Proportion of water service availability indicator at public healthcare facilities in Addis Ababa city, Ethiopia 2022.

| Water service indicators at HCF | Proportion by HCF |  |  |
| --- | --- | --- | --- |
|  | Hospital<br>N(%) | Health center<br>N(%) | Total<br>N(%) |
| Water availability during survey (N=86) |  |  |  |
| ▪ yes | 11(100%) | 63(84%) | 74(86%) |
| ▪ no | 0(0%) | 12(16%) | 12(14%) |
| Water discontinuity (N=86) |  |  |  |
| ▪ yes | 7(63.6%) | 57(76%) | 64(74.4%) |
| ▪ No | 4(36.6%) | 18(24%) | 22(25.6%) |
| Frequency of discontinuity (N=64) |  |  |  |
| ▪ Frequently per day | 2(28.6%) | 27(47.4%) | 29(45.3%) |
| ▪ rarely per day | 5(71.4%) | 30(52.6%) | 35(54.7%) |
| Alternative water source (N=86) |  |  |  |
| ▪ yes | 6(54.5%) | 4(5.3%) | 10(11.6%) |
| ▪ no | 5(45.5%) | 71(94.7%) | 76(88.4%) |
| Piped water at OPD (N=86) |  |  |  |
| ▪ yes | 8(72.7%) | 39(52%) | 47(54.7%) |
| ▪ No | 3(27.3%) | 36(48%) | 39(45.3%) |

**HCF:** Healthcare Facility

### Sanitation service availability

All healthcare facilities, 86(100%) had limited sanitation services and no one the healthcare facilities had access to basic sanitation services. Of the total, 49(57%) healthcare facilities had usable toilets. Only 4(4.7%) of healthcare facilities and 17(19.8%) of healthcare facilities had access to menstrual hygiene toilets and toilets accessible for users with limited mobility respectively (Table 2).

**Table 2:** Sanitation service indicator at public healthcare facility in Addis Ababa city, Ethiopia 2022.

| Sanitation services indicator | Proportion by HCF type |  |  |
| --- | --- | --- | --- |
|  | Hospital<br>N (%) | Health<br>center<br>N (%) | Total<br>N (%) |
| Types of Toilet |  |  |  |
| ▪ flush/pour flush to sewer system | 6(54.5%) | 2(2.7%) | 8(9.3%) |
| ▪ flush/pour flush to septic tank or pit | 5(45.5%) | 71(94.7%) | 76(88.4%) |
| ▪ Pit latrine with slab | 0(0%) | 2(2.7%) | 2(2.3%) |
| Usable toilet |  |  |  |
| ▪ yes | 7(63.3%) | 42(56%) | 49(57%) |
| ▪ no | 4(36.4%) | 33(44%) | 37(43%) |
| Staff toilet |  |  |  |
| ▪ yes | 9(81.8%) | 62(82.7%) | 71(82.6%) |
| ▪ no | 2(18.2%) | 13(17.3%) | 15(17.4%) |
| Menstrual hygiene toilet |  |  |  |
| ▪ yes | 0(0%) | 4(5.3%) | 4(4.6%) |
| ▪ no | 11(100%) | 71(94.7%) | 82(95.4%) |
| Toilet for limited mobility |  |  |  |
| ▪ yes | 0(0%) | 17(22.7%) | 17(19.8%) |
| ▪ no | 11(100%) | 58(77.3%) | 69(80.2%) |
| Excreta/waste-water disposal |  |  |  |
| ▪ Sewerage system | 6(54.5%) | 4(5.3%) | 10(11.6%) |
| ▪ Septic tank | 5(45.5%) | 71(94.7%) | 76(88.4%) |
| Urinal service for male |  |  |  |
| ▪ yes | 2(12.8%) | 9(12%) | 11(12.8%) |
| ▪ no | 9(81.8%) | 66(88%) | 75(87.2%) |

### Hand hygiene service availability

Out of 86 observed healthcare institutions, only 7(8.1%) institutions, one hospital, and six health centers, had access to basic hand hygiene services, while three healthcare facilities (3.5%), one hospital and two health centers, had not to hand hygiene service both at point of care and within 5 meters of the toilet. Majority of healthcare institution had limited hand hygiene service, meaning that they had not to hand hygiene service both at the point of care and nearby the toilet (Figure 3).

**Figure 3:**
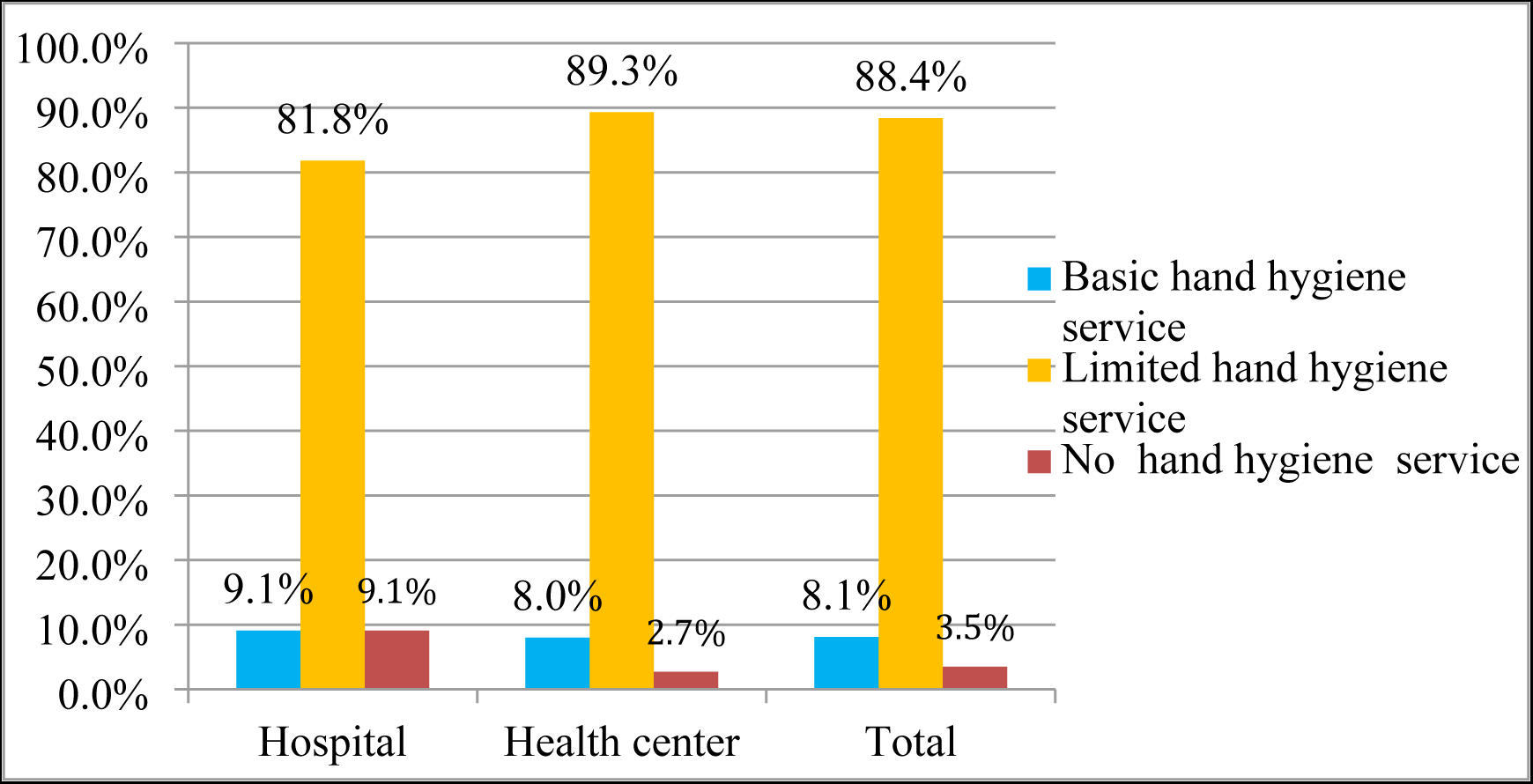
Hand hygiene service availability at public healthcare facilities in Addis Ababa city, Ethiopia 2022.

Of all, 59(68.6%) of healthcare facilities had functional hand hygiene facilities (either with water and soap or alcohol hand rub) at point of care. However, 11(12.8%) of healthcare facilities had not to hand hygiene service at the point of care.(Table 3).

**Table 3:** Proportion of Hand Hygiene service status at public healthcare facility in Addis Ababa city, Ethiopia 2022.

| Hand hygiene services indicator | The proportion by HCF type |  |  |
| --- | --- | --- | --- |
|  | Hospital<br>N(%) | Health<br>center<br>N(%) | Total<br>N(%) |
| Hand hygiene facility at point of care<br>(N=86) |  |  |  |
| ▪ Yes functional hand hygiene with water and soap | 2(18.2%) | 10(13.3%) | 12(14 %) |
| ▪ Yes Alcohol based hand rub(ABHR) | 6(54.5%) | 41(54.7%) | 47(54.6%) |
| ▪ Yes but it lacks water and/soap | 2(18.2%) | 14(18.7%) | 16(18.6%) |
| ▪ No hand hygiene service | 1(9.1%) | 10(13.3%) | 11(12.8%) |
| Hand hygiene facility within 5 meter of the toilet (N=86) |  |  |  |
| ▪ Yes functional hand hygiene | 1(9.1%) | 9(12%) | 10(11.6%) |
| ▪ Yes but it lacks water and/soap | 8(72.7%) | 54(72%) | 62(72.1%) |
| ▪ No hand hygiene service | 2(18.2%) | 12(16%) | 14(16.3%) |
| Hand hygiene Promotion material on washing facility (N=72) |  |  |  |
| ▪ yes | 1(11.1%) | 4(6.3%) | 5(6.9%) |
| ▪ no | 8(88.9%) | 59(93.7%) | 67(93.1%) |
| Hand hygiene facility accessible to all users<br>(N=72) |  |  |  |
| ▪ yes | 1(11.1%) | 4(6.3%) | 5(6.9%) |
| ▪ no | 8(88.9%) | 59(93.7%) | 67(93.1%) |

### Healthcare waste management service availability

Out of 86, healthcare facilities studied 25(29%) had not to waste management service at all. Unfortunately, only one (1.2%) facility had basic healthcare waste management service. More than two thirds 60(69.8%) of Public healthcare facilities in the city of Addis Ababa had limited waste management service (Figure 4).

**Figure 4:**
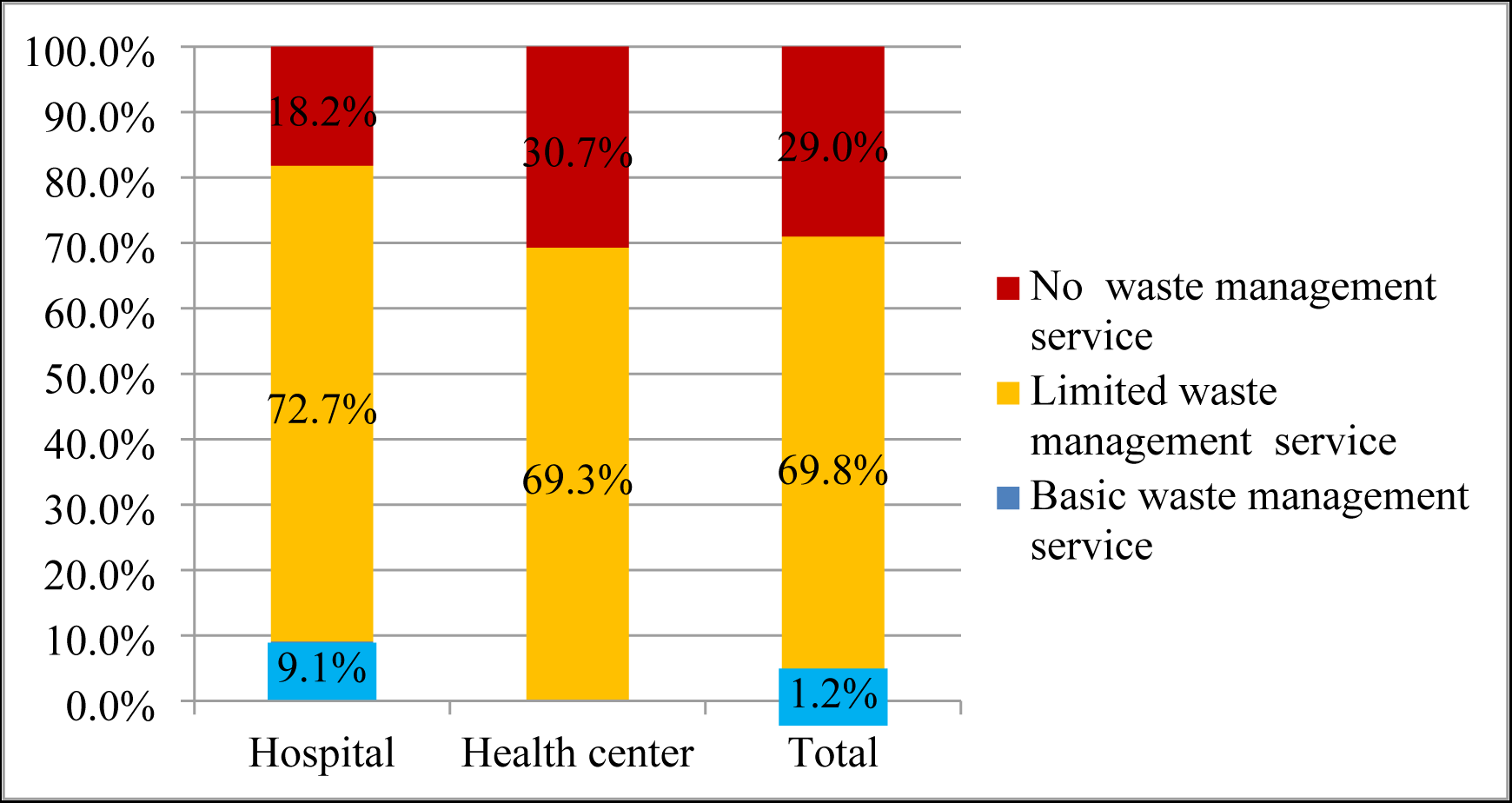
Healthcare waste management service availability at public healthcare facility in Addis Ababa city, Ethiopia 2022.

Of all healthcare facilities, 25(29%) of facilities had not to waste segregation bins in the outpatient department; only one hospital safely segregated medical waste in three labelled bins. Seventy-nine (91.8%) healthcare facilities and 80(93%) healthcare facilities used brick-type incinerators for the disposal of infectious waste and sharp waste respectively. A protected pit was used by all healthcare facilities to dispose placenta and pathological waste. All healthcare facilities dispose of pharmaceutical waste at the national waste collection point in collaboration with Food and Drug Administration Authority (FDA), Ministry of Health, and Addis Ababa Regional Health Bureau (Table 4).

**Table 4:** Proportion of healthcare waste management service status at public healthcare facilities in Addis Ababa city, Ethiopia, 2022.

| Waste management service indicators | Proportion by HCF type |  |  |
| --- | --- | --- | --- |
|  | Hospital<br>N(%) | Health<br>center N(%) | Total<br>N(%) |
| Medical waste segregation |  |  |  |
| ▪ Waste segregation meet the standard | 1(9.1%) | 0(0%) | 1(1.2 %) |
| ▪ Segregation and bins aren't meet the standard | 8(72.7%) | 52(69.3%) | 60(69.8%) |
| ▪ segregation bins are not present | 2(18.2%) | 23(30.7%) | 25(29%) |
| Disposal of infectious waste |  |  |  |
| ▪ incinerator(two-chamber,850-1000) | 2(18.2%) | 0(0%) | 2(2.3%) |
| ▪ incinerator(brick type) | 7(63.6%) | 72(96%) | 79(91.8%) |
| ▪ burning in the protected pit | 0(0%) | 3(4%) | 3(3.5%) |
| ▪ Collected for disposal off-site | 2(18.2%) | 0(0%) | 2(2.3%) |
| Disposal of sharp waste |  |  |  |
| ▪ incinerator(two-chamber,850-1000) | 2(18.2%) | 0(0%) | 2(2.3%) |
| ▪ incinerator(brick type) | 7(63.6%) | 73(97.3%) | 80(93%) |
| ▪ burning in protected pit | 0(0%) | 2(2.7%) | 2(2.3%) |
| ▪ Collected for disposal off-site | 2(18.2%) | 0(0%) | 2(2.3%) |

### Environmental cleaning service availability

Only 2(2.3%) healthcare facilities had basic environmental cleaning services, while the rest 84(97.7%) healthcare facilities had limited service (Figure 5).

**Figure 5:**
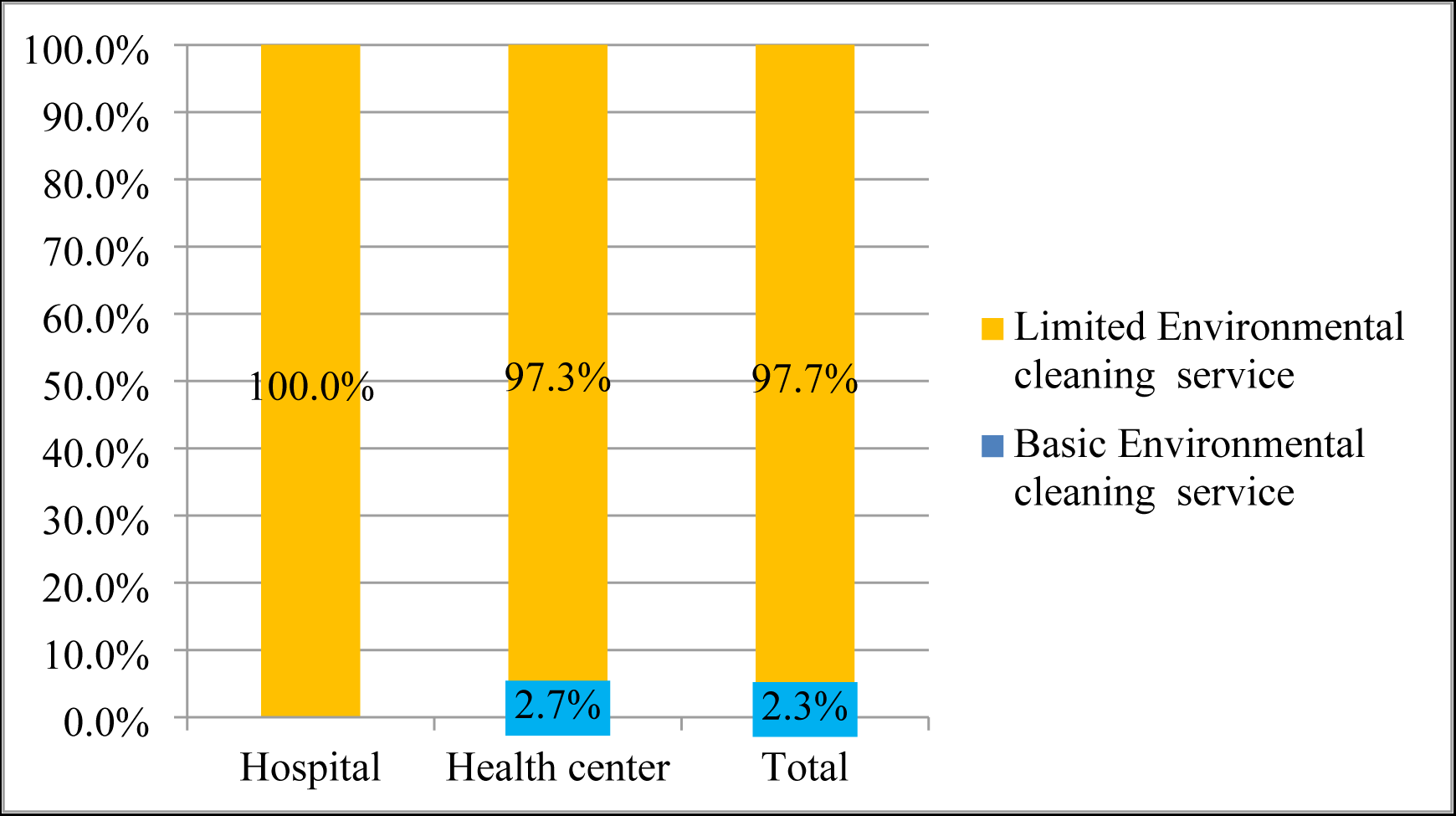
Environmental cleaning service availability in Addis Ababa public health care facility, Ethiopia 2022.

About one quarter 22(25.6%) of health care facilities had cleaning protocols for all cleaning services with cleaning schedules, and only 2(2.7%) health centers trained all staff related to cleaning services and standards. Furthermore, only 7(8.2%) of healthcare facilities had cleaning supplies including cleaning equipment and detergent in the patient care area (Table 5).

**Table 5:** Proportion of environmental cleaning service status of public health care facility in Addis Ababa city, Ethiopia, 2022.

| Environmental cleaning service indicators | Proportion by HCF type |  |  |
| --- | --- | --- | --- |
|  | Hospital<br>N(%) | Health center<br>N(%) | Total<br>N(%) |
| Cleaning protocol availability |  |  |  |
| ▪ Yes for all cleaning service | 9(81.8%) | 13(17.3%) | 22(25.6 %) |
| ▪ Yes, but not for all cleaning service | 2(18.2%) | 62(82.7%) | 64(74.4%) |
| Training for responsible staff |  |  |  |
| ▪ Yes for all given | 0(0%) | 2(2.7%) | 2(2.3%) |
| ▪ Some staff had trained | 11(100%) | 70(93.3%) | 81(94.2%) |
| ▪ No one is trained | 0(0%) | 3(4%) | 3(3.5%) |
| Cleaned floor, table & unpleasant smell |  |  |  |
| ▪ yes | 11(100%) | 64(85.3%) | 75(87.2%) |
| ▪ no | 0(0%) | 11(14.7%) | 11(12.8%) |
| Cleaning supplies in the area of the outpatient room |  |  |  |
| ▪ Yes present | 6(54.5%) | 1(1.2%) | 7(8.2%) |
| ▪ Not present | 5(45.5%) | 74(86%) | 79(91.8%) |

### Healthcare WASH service barriers

#### Socio-demographic characteristics of key informants

For the qualitative interview, 16 key informants have participated. Of these, 11 were recruited from hospitals, 4 were from health center, and one was from Addis Ababa Regional health bureau. Among the participants, 12(75%) were male. The participants were between the age of 26 and 58 with a mean age of 35.25 ± 7.77 years had 3 to 39 years of working experience with a mean of 10.94 ± 9.23 years. Thirteen and three participants had bachelor’s degrees and master’s degrees educational status, respectively. Of these, 11(68.7%) were working as infection prevention and control officers, 4(25%) participants were working as medical directors, and 1(6.3%) participant working as WASH program officer, respectively. The average time for the interview was 26.16 minutes.

#### Healthcare WASH service barriers

The qualitative parts of the study consist of five main themes and 12 subthemes of barrier, to providing basic healthcare WASH services, raised by the participants.

**Built Environment and Necessity:** the most frequently quoted challenges by participants; it describes limited availability, poor design, none functioning, poor maintenance service of healthcare WASH infrastructure and shortage of necessities like water affects healthcare WASH service provision. This theme emerged from four subthemes, non-availability (inadequate) of basic WASH infrastructure; lack of maintenance service; poor design and layout of facility and WASH infrastructures; and shortage of necessities.

**Resource availability and allocation:** in this theme, the availability of resource and allocation of it affects the status of WASH services. Participants mention that the inadequate procurements of supplies, extended bid system, high maintenance cost, and inadequate (lack) budget allocation for WASH impeded healthcare WASH service. The subthemes under this theme were inadequate (lack) budget allocation and shortage of supplies.

**Stakeholder participation and Leadership commitment:** this theme describes the lack of supervision and commitments of facility managers, and responsible governmental institutions, and the lack of partner organization participation are among the barriers to providing basic healthcare WASH service. Emerged subthemes are the lack of administrative support and leadership; and lack of partner organizations participation.

**Individual level barrier:** this theme describes individual level awareness, attitude, and behaviour, able to improve or retard the level of healthcare WASH service. The emerged subthemes under this main theme are lack of training and awareness; and poor attitude and behaviour.

**Legal issue barrier:** this theme describes the role of framework and guidelines on healthcare WASH service. Participants said the poor status of healthcare WASH service is attributed to unavailability of framework supported by healthcare WASH program documents and guidelines for a year. Emerged themes are the lack of program-supported framework, and the lack of healthcare WASH guidelines.

### 1. Built environment and Necessity related barrier

This main theme describes the most frequented barriers related to hardware infrastructure and the necessity to provide basic healthcare WASH service as mentioned by participants. Most of the time, the available WASH facilities were not fully functioning because of a lack of timely maintenance and renovation services. Another barrier related to the build environment is the design of the building and WASH hardware; each service point of the facility had not WAHS system because of the poor design of the facility. In addition, participants state that shortage of water is a critical challenge for healthcare WASH services.

#### A. Non-availability of basic WASH infrastructures

The participants described healthcare facilities are not doing WASH services as per the standard. The minimum requirement of basic WASH infrastructure has not been available and it was not in line with client flow and as per the standard. Senior program officer participant stated that;

> *“Availability of built facility (hardware) is a basic challenge; you know…all the facilities were not constructed considering adequacy and the inclusive WASH issues that create a significant challenge in the facility itself to address the issue right now”. (KI 11)*

Participants noted that the construction of WASH facility at healthcare setting was not considered users continence and inclusive at the first beginning to address the issue of WASH for all; which was inadequate, and lack of user-sensitive toilets impact the usability of WASH service and compromise the needs of clients and facility staffs at all. The female deputy medical director of the health center stated that;

> *“…we do have a narrow outpatient room (small in surface area by m2) without hand washing facility. We do have also small number of toilets; the users and number of toilets are not much, we do not have toilets for menstrual hygiene service, also we do not have toilets for limited mobility customers including staff. Personally I fear what will happen to these existing WASH services if cholera occurred at this time”. (KI 14)*

Despite participants recognizing the significance of WASH service availability at each point of care and critical area, there was a variation of availability among service points. A medical director from the health center stated that

> *“….some rooms have not to WASH service equipment (hardware facility). As you know every service provider at the point of care should wash their hand after a visit or counselling each patient but most of our service point rooms had not to hand washing facility”. (KI 12).*

The number of WASH facilities and users was not balanced, and participants were stressed on the ratio of expected users to available hardware should be critically designed and planned at the time of healthcare facility construction. An infection prevention officer (IPC) from one hospital described;

> *“…One of the challenges is WASH infrastructure availability; for example, in our hospital, there are no built washing sinks (hand washing) at each point of care or service point. And also the hospitals have inadequate toilets”.(KI 04)*

#### B. Lack of maintenance service of WASH infrastructures

The other barrier described by participants was the lack of a functional WASH facility and poor maintenance services of it. Not all WASH facilities were fully functioning and ready to use because of a lack of active maintenance and renovation services. A senior IPC officer from one hospital stated;

> *“….from my experience, it is common to see broken or non-function toilets, sinks, and other WASH services hardware in this hospital, which are not maintained timely the same time. There is a maintained inability to make the service fully (100%) functional in a timely and sustainable manner”. (KI 02)*

Most of the healthcare facility had not sanitary maintenance workers and electricians at the time of the survey and the state of sustained functionality of WASH infrastructure are questionable; due to that lack of actively tracking and maintenance service was challenging for the healthcare facilities. IPC officer from one hospital described;

> *“…Lack of sustainability issues for the existing WASH facilities is a challenge in our hospital; what I have observed in our facility is there is no culture of prevention which means there is no active preventive maintenance and repair service for WASH systems rather corrective one”. (KI 08).*

#### C. Poor design of WASH infrastructure

Another barrier related to the build environment is the design and layout of the building and WASH hardware; participants explained that how the design of the building and installation of WASH hardware substantially enhance or retarded healthcare WASH services. The program officer from Regional Health Bureau explained that;

> *”…The major bottlenecks are related to the design of buildings, if you experience the available health centers in the city the toilets and hand washing facilities are not functional which is mainly resulted from the poor design and installation of WASH facilities”.(KI 11)*

As stated by participants, each service point of the facility had not all WAHS system because of the poor design of the facility; even if the hardware is available, it might not be fully functional because some of the facilities (hospitals) were built many years ago and they need to be renovated, and some of the hardware has not been properly installed. The health center medical director stated;

> *“…If we observe the service delivery areas, they don’t have water service; because water pipe is not installed towards many of the service areas in the first beginning, and also the rooms (service point) had not installed hand washing facility. Sometimes, I thought like…. the building was not built for healthcare facility at the first beginning”. (KI 16)*

#### D. Shortage of necessity

In all healthcare facilities, shortage of water was found to be challenging for healthcare WASH service provision; even if the hardware is available, shortage of water was mentioned as a challenge to make fully functional all other WASH services across the building. Another deputy medical director from the health center explained;

> *“…we are sharing water with the community, we do not access water independently and water is interrupted for three* (*3*) *or four*(*4*) *days…even we do have a limited number of tankers as a backup of water to serve us a couple of days and it will run away at the end”. (KI 13)*

### 2. Resource availability and allocation related barriers

In the healthcare setting, the availability and allocation of adequate resources have played a significant role in the hardware and software components of WASH service. The participant mentioned that the availability and practice of healthcare WASH service was affected by inadequate budget allocation for WASH, inadequate procurements of supplies, extended bid system, and high maintenance cost; An IPC officer from the hospital highlighted that;

> *“The challenge is no separate budget for the IPC and WASH service and there is frequently a shortage of WASH-related supplies. We did not ask them why supplies run out frequently and why purchased in a timely because there is no separate budget code assigned to WASH activities, even not for IPC”. (KI 07).*

Participants said that not only a shortage of resources but also unable to use dedicated budget effectively was found to be a barrier; poor supply procurement habit of administration and extended or delayed bid system was challenging the healthcare WASH service. One of IPC officers from the hospital supported it;

> *“The WASH service is not available similarly in each room due to a shortage of supplies. For instance, lack of segregation of biohazard wastes; the purchasing process of the government (facility administration) is very lagging. Lack of sharp waste collecting bins (safety box) is also one of our problems; we use safety boxes prepared from normal cartons which are not as per the standard. This is available on the market system, but there is purchasing delay”. (KI 02).*

### 3. Stakeholder participation and Leadership

This theme describes the lack of supervision and commitments of facility managers and responsible governmental institutions, and the lack of partner organization participation are among the barriers that hampering basic healthcare WASH access. Emerged subthemes are inadequate administrative support and poor leadership, and lack of partner organization participation.

#### A. lack of administrative support and leadership

A great challenge is the lack of actively engaging in the implementation of WASH activities. The participants’ explanation was agreed that the improvement of healthcare WASH services needs the engagement and support of governmental administration and committed health sector and facility leadership. This is the problem of most of the facilities at the time of the survey challenging back the healthcare WASH service, with a little bit of variation, the management staff either from the facility or higher offices did not have commitments to share and fix the challenges. One of the hospital IPC officers stated that;

“One of the major challenges is lack of senior management engagement, they do not consider the WASH activities as the major one rather considering it as additional and/or auxiliary activity and task aside to the clinical service. For instance, they did not allocate budget for the supplies and maintenance service, they did not conduct routine support for WASH activities” (KI 07)

The problem was magnified at the health center level, which denied them from seeking financial and technical support from higher health sector offices like sub-cities and regional health bureaus. The health center depute medical director described that;

> *“…We do not have supportive supervision and follow-up from sub-city or any higher office regarding Health center WASH services. Since in my career in this facility, no one is visit here until now, to support and supervise WASH service specifically unlike other clinical service” (KI 14).*

Unlike the clinical service, WASH service was not emphasized by the Regional Heath Bureau and Ministry of health starting from the design and construction of the health facility to the implementation of the WASH program, participants described how this made challenging for the existing healthcare WASH service. The program officer supported this statement;

> *“The major problem is during the construction of buildings when the health facilities are constructed, it is based on the interest of engineer’s taken the contract. Health care professionals and WASH experts are not participating in the design of buildings”.(KI 11)*

Health sector governance has not given attention to allocating WASH practitioners in the healthcare facility, particularly at health centers. Participants described that challenging of limited or shortage of technical personnel assigned for effective implementation of healthcare WASH service, unavailability of environmental health and hygiene experts in the health facilities (87% of healthcare facilities) is affecting the system which resulted from lack of emphasis given for healthcare WASH services. The WASH program officer highlighted that;

> *“Most of the health facilities use health officers or other professionals at IP (infection prevention) focal persons but no environmental health professional is deployed for the position. So, the challenge is related to the shortage of environmental health professionals”. (KI 11)*

#### B. lack of partner organization participation

Some of participant rose that, the other challenge to limited healthcare WASH service is due to the absence of partner organization participation in healthcare WASH project. The participant’s opinion described that engagement of partner organizations would improve and sustain basic healthcare WASH service yet that did not happen. The unavailability of agreement between partner organizations and healthcare facility administering body is the unmet opportunity for healthcare WASH services. The IPC officer highlighted this;

> *“…I think lack of engagement of partner organizations like NGOs…..or governmental stakeholders working on it (WASH), is just one of the enabling environments for limited services of healthcare WASH. Renovation of hospitals like waste management technology including incinerator, waste drainage system could be upgraded and sustained by external stockholder yet not that happened on the ground”. (KI 04)*

Although partner organizations are participating in many clinical service programs in most of the surveyed hospitals, the participants indicated that the WASH service was overlooked in the health settings. Participants believed that the constraints of resource and technology limitations of the facility could be overcome by engagements of partner organizations, as highlighted by the deputy medical director;

> *“We need support from partner organizations because they could be the source of funds, there may do have alternative technology and improve all over the status of WASH both in hardware and capacity building aspects”.(KI 14)*

### 4. Individual level barriers

This theme describes individual level awareness; attitude and behaviour can improve or retard the level of healthcare WASH service. The emerged subthemes under this main theme are lack of training and awareness; and attitude and behaviour.

#### A. Lack of Training and Awareness

The training was given to healthcare professionals including cleaners across healthcare facilities. However, participants pointed out how low levels of awareness and inadequate training on healthcare WASH among staff and allied workers were challenging to healthcare WASH service, leading to the poor practice of it. The IPC officer said that;

> *“Even though the hospital is continuous professional development (CPD) center, healthcare workers did not take adequate training; They do have a low level of awareness on the importance of healthcare WASH which leads to poor adherence to WASH service practice during healthcare attachment particularly poor hand hygiene compliance and waste management practice”(KI 03)*

#### B. Poor Attitude and Behaviour

Participants said that both professionals and management did not feel that WASH activities are the responsibility of all other staff beyond designated IPC officers and cleaners; They considered WASH is just an IPC, and they did not want to take a role in the improvements of healthcare WASH service. IPC officer from one hospital noted;

> *“One of the first challenges is attitude problem on IPC and WASH because IPC is not individual responsibility its responsibility of all staff, patients and management bodies; however, some staffs even management bodies perceived that WASH is just an IPC and it is only the duty of the focal person or IPC officers”. (KI 04)*

There was also professional negligence and ignorance towards the importance of healthcare WASH service at the point of care; participants pointed out that some professionals were not adhering to the practice of cleaning protocols. An IPC officer from one hospital described that;

> *“There are a physician and other health professionals who perceived that ICP is just bout hand washing; so that they do not mind about WASH activities like waste segregation at point of care, cleaning of point of care and the like”.(KI 02)*

Similarly, participants noted that negligence of proper waste segregation among clients was found one of the challenges to healthcare WASH services, as spoken by the IPC officer;

> *“Waste segregation practice is very poor among caregiver of the patient and health professionals have poor practice in waste segregation. For example, they damp gloves inappropriately but mostly waste segregation problem raised from the client, caregiver and visitors and we try to face the problem by giving health education”. (KI 05)*

### 5. Legal issue towards healthcare WASH

Participants believed that the existing poor healthcare WASH service was attributed to unavailability of a holistic WASH framework supported by healthcare WASH guidelines. Unavailability of healthcare associated infection surveillance systems, and unavailability of guidelines in healthcare settings was a challenge to WASH service provision. An IPC officer from the hospital stated that;

> *“There is no working healthcare WASH framework unlike other clinical services given in the hospital. There is no surveillance system and tracking staff in the hospital that conduct tracking of healthcare associated infections”. (KI 03)*

Among the surveyed healthcare facility not more than 4 facilities started to apply and practice new healthcare WASH guideline; hence unavailability of specific healthcare WASH guideline for a year was also a challenge as participants raised. An IPC officer explained it;

> *“There is no specific WASH document or officer. WASH activities are overlooked in healthcare setting; there is IPC guideline and IPC focal officer who runs WASH integrated with the IPC department. Healthcare facilities had no independent guideline and professional staff responsible for primarily to WASH service”. (KI 10)*

## Discussion

The findings of this mixed study provide the current status of healthcare WASH service and the contemporary opinion of participants (healthcare professionals) experience on healthcare WASH service barriers in public healthcare facilities of Addis Ababa City. In this study, we found that no one healthcare facility had basic access to all WASH services. The independent WASH domain analysis showed that about 86% of healthcare facilities had basic water access, 100% had limited sanitation access, 88.4 % had limited hand hygiene service, 69.8% had limited healthcare waste management service, and 97.7% had limited environmental cleaning service. Compared with the findings of other studies, overall access to basic healthcare WASH service in our study was lower than from the study conducted on urban healthcare facilities in Uganda (12.12%) (21), and a study conducted on rural healthcare facilities in sub-Saharan Africa countries e.g: Zambia (21%), Kenya (30%), Uganda (30%) and Ruanda (50%) (22). The limited access to healthcare WASH service might be due to unavailability of healthcare facility WASH standards for a year, and the lack of committed leadership from the side of the government. The other possible explanation could be due to unavailability of adequate resources distribution across healthcare facilities for the WASH services, building, and maintenance of infrastructure.

To our findings, healthcare facilities are facing challenges in providing basic access to WASH service. In this study, the finding was different from the national level of WASH service reported by the WHO and UNICEF joint monitoring healthcare WASH baseline report. The basic access to water service (86%) was higher than from national level access (30%) (18), and the worldwide level of basic water service (78%) (6). However, the available basic sanitation service(0%) and basic hand hygiene service (8.1%) in our study were lower than from national level of access 59% and 52% respectively by 2016 (18), and from the study conducted in the north western part of Ethiopia, 21.4% of facility had basic access to hand washing facility (23). The higher level of basic water service access might be attributed to the study area conducted in urban setting, which is the capital city of Ethiopia with better investment to access improved water sources as compared to studies conducted in urban and rural parts of the country. The lower level of basic hand hygiene service could be due to the limited availability of financial resources to facilitate supplies and maintenance service of hand hygiene facilities and the lack of timely repair system across healthcare facilities.

In this study, all healthcare facilities had limited sanitation service which is lower than in sub-Sahara African countries, 13% of healthcare facilities had basic sanitation service by 2022 (6); only 5% and 20% of healthcare facilities had access to a toilet for menstrual hygiene and limited mobility clients respectively; which means that more than 80% of public healthcare facilities in Addis Ababa were not user sensitive. These are more likely to affect the dignity and privacy of users. Unavailability of insufficient gender-sensitive sanitation facilities, the menstrual hygiene toilets, was also shown in studies conducted in Uganda and Zimbabwe (21, 24), the proportion of sanitation facilities for menstrual use and disabled user remains poor. This could be due to that the essence of WASH for all might not be given emphasis and the design of the facility not considering user sensitive sanitation facility.

This study identified healthcare WASH service barriers, participants provided detailed descriptions of their healthcare facility WASH barriers, and the identified themes explained the limited access to healthcare WASH service findings. We found that built environments of WASH related barriers; lack of resource availability and allocation; partner organization and leadership barrier; individual level barriers and legal related barriers were the identified barriers to healthcare WASH service provision in the study area.

In the context of poor healthcare WASH services, adverse events including healthcare acquired infection and risk of AMR significantly affect the healthcare system (23). In our study, built environments of WASH infrastructure was found to be the most commonly cited barrier to WASH services provision in all healthcare facility. The unavailability of the built facility, lack of maintenance service, and poor design of the facility were among the significant barriers that were affecting the healthcare WASH services. The findings were found to be consistent with studies conducted in Kenya and Ethiopia, and the JMP baseline report (25–27). The design and availability of built WASH facilities were the most prevalent challenges; facilities were not built considering adequacy for users, and facilities were not designed at every point of care and did not consider special needs in the healthcare settings; maintenance service was not also given attention. These all affect the availability and practice of WASH services in the healthcare settings to the prevention of healthcare acquired infection and COVID-19 disease (28).

In the absence of an adequate and user friendly built sanitation service, an estimated risk of transmission of hospital acquired infection by contaminated environment contributed to 30-50% (29). Healthcare facilities were not in the position to improve the dignity and privacy of patients, staff, women and people with limited mobility regarding WASH services. The descriptive parts of our findings indicated that 95% of facilities had not to gender sensitive WASH service which makes to feel discomfort among menstruating girls and women due to issues of privacy and dignity (30); 80% of facilities had no a toilet for limited mobility users, this physical barrier kept away the disable users from using the toilet in the facility (31); 17% of healthcare facilities had not staff toilet, and 87.2% of the facility had not urinal service for men nearby the toilet. Due to that existing built sanitation facilities in the study area could have the significant contribution to poor quality of care and increase the risk of getting an infection (29,32,33).

Lack of functional hygiene facility at healthcare setting has the potential to increase the risk of healthcare acquired infection at the point of care and within the facility compound (29, 33). In this study, due to physical, financial, and leadership related barriers raised by participants, 31.2% and 88.4% of healthcare facility had not functional hand hygiene facilities at the point of care and nearby the toilet respectively, either it has not to hand hygiene services at all, lacks water and/or soap at time of survey or it lacks maintenance services; and 93% of hand washing facility was not accessible to users with special need.

Similarly mixed method evaluation in part of Ethiopia (23), lack of functional hand hygiene facility and lack of washing materials at the point of care and nearby the toilet was impeding factors indicating that quality of care and patient safety was compromised across all healthcare facility, which makes that the risk of catching with COVID-19, monkey box, Ebola and any else among patients and caregivers are high (32, 34).

Shortage of water was found to be one of the main barriers in most the healthcare facilities surveyed; Interruption of water availability affects the function of WASH service. This finding was also highlighted from studies conducted in Uganda and India (21, 30). In our finding, facilities were suffering from a shortage of water; three-quarters of healthcare facility were faced water discontinuity previous to the survey due to that healthcare facilities were accessing water through shifting (*fereka*) program with the community; of which nearly half(45.3%) of the facility was experienced daily water interruption. In the healthcare setting, having only a built facility is not good enough to prevent infection; all WASH domains must work synergistically; in the absence of water service toilet and hand washing facility would nor functional, clients also would not use the WASH facility and exposed to infection and which has led to developing negative experience and frustration that will deny them seeking care and loss of trust on the future (9, 27).

The other most frequently cited barrier in our finding was inadequate resource availability impeding the availability of basic healthcare WASH service in the facilities. This was in line with studies conducted in Ethiopia and Kenya about resource barriers to health care WASH (13, 25). The challenges in the prevention of infection have doubled in countries with the limited resources at the time of covid-19 pandemic (35). To avail all the required healthcare WASH facilities, to provide adequate and timely training for healthcare staff, and timely maintenance of services, basic financial resources and supplies are required and government should allocate adequate budget and materials. Similar to the qualitative study conducted in Malawi (23), a shortage of WASH supplies affect the availability of WASH services and practice in the study area.

Shortage of supplies (materials and equipment) for healthcare WASH service was found to be a significant barrier that hampering healthcare professionals, janitors, and clients to practice hygienic behaviours and environmental cleaning services. Shortage of WASH supplies causes to miss handling and management of healthcare waste, poor cleaning services, and poor hand hygiene practices across healthcare facilities, these could be due to the unavailability of budget and poor management of supplies which makes that care provider, patients, and cleaners vulnerable to infection and poor quality of care in the facilities. This challenge similarly reported from studies conducted in Ethiopia (10, 27), highlighted the significance of adequate WASH service supplies to safeguard healthcare providers and to enhance the quality of care. Therefore, a shortage of supplies do not build trust in the working environment and affects prevention practices (34, 36); this implies that patient care practice is compromised that able to facilitate the chance of getting healthcare acquired infection or any emerging disease among patients, caregivers, waste handlers, and visitors across the healthcare facilities (37, 38). These all could be attributed to a lack of and/or inadequate budget allocation for WASH services and a delayed bid system to purchase WASH materials and equipment.

Our finding showed that nearly one-third of the facilities in the study area had no healthcare waste segregation bins at the point of care, and two third of facilities had limited waste segregation services. Later shortage of waste segregation bines including safety boxes at the point of care leads to the poor practice of waste management and exposure to occupational hazards in the facility (38, 39).

The other major barriers reported in this study were the lack of committed leadership and stakeholder participation. To the improvement of healthcare WASH service engagement and support of governmental administration, and committed health sector and facility leadership are worthy issues. Studies reported the implementation of basic WASH services in the healthcare setting is affected by the absence of committed leadership, lack of timely and on-going monitoring system for implementation and maintenance, cleaning and waste management system; and the absence of a range of stakeholders capable of influencing or responsible for the provision of sustainable basic healthcare WASH services (13, 25).

In this study, the most challenging in most healthcare facilities was, senior management staff, either from the facility or higher offices, were not taking a role in WASH service improvement in a good way. They did not allocate enough budget and technical staff for WASH service; they were not actively to engage in the design and construction of the facility; they were not providing timely technical assistant to IPC team to improve healthcare WASH service. Similarly, studies reported the degree of commitment and engagement of government and senior facility managers to take an action to improve healthcare WASH services was significantly attributed to the variation in WASH services availability in the healthcare facilities (8,21,25).

Evidence showed that the lack of integration of WASH service with other key national programs, and the lack of participation of private or none profit organizations in healthcare WASH projects were affecting the status and sustainability of healthcare WASH services. To solve the financial and technical barriers, integration of programs and agreements with partner organizations could be an alternative solution for healthcare facilities with the limited resources (13, 29). In this study, the lack of engagement of partner organizations in healthcare WASH, and excluding the city from the One-WASH program was mentioned as barriers to WASH service provision. This might be due to that the lack of an effort to institute functional multi-sectorial coordination and technical working force to healthcare WASH service by the ministry of health and regional health bureau.

This study found that individual level barrier was affecting healthcare WASH service; participants noted that lack of awareness, lack of training, poor attitude and negligence among care providers and janitors, and lack of awareness and inattention of healthcare attendants in the study area were the barriers for healthcare WASH service practice. The study found to be consistent with other findings in Ethiopia, inadequate training on WASH and IPC, low awareness of janitors and healthcare workers, and visitors were challenged to improving and manage healthcare WASH service in the facility (27,37,40).

We found that healthcare WASH service was suffering from the absence of healthcare WASH framework and guidelines. Participants in this study noted out the absence of compressive healthcare WASH service framework was attributed to limited healthcare WASH service in the facilities; healthcare WASH framework provides a clear road map for healthcare facility including targets to be achieved, healthcare infection surveillance system, monitoring tool and integration of it with the other clinical services. The other barrier raised by participants was the lack of healthcare WASH guidelines for a year. The absence of updated and inclusive guidelines focusing on healthcare WASH was significantly affecting facilities. Healthcare WASH facilities were not adequate, inclusive, and user friendly as a result of the absence of updated guidelines. The finding was in line with studies (8,13,29), gaps in the national level healthcare WASH framework and guidelines were the barriers to improving healthcare WASH service.

## Strengths and limitations of the study

As a limitation, since the key informants were selected purposely generalization of healthcare WASH service barriers to the larger population cannot be made. As the strength, the study used a mixed method design by observing the WASH facilities and involving the key informant interview to have a complete understanding of the availability of healthcare WASH services and the barriers to WASH services provision. Using the new healthcare WASH service assessment tool, including details of WASH domains, accurately revealed the status of WASH services in healthcare facilities.

## The implication of the Study

This study is the first in kind to assess the availability of healthcare WASH services based on the JMP service ladder, and to describe the experience of IPC focal officers, medical directors, and program officer about the barriers to healthcare WASH services provision in Ethiopia. Despite improvements of WASH related hygienic behaviour at the time of the COVID-19 pandemic, the study revealed inadequate WASH services and multiple challenges across healthcare facilities have negative implications on the prevention and control measure of the COVID-19 disease, healthcare acquired infection and increase risk of AMR, and emerging future pandemics.

In the absence of significant improvement in basic service of healthcare WASH services, COVID-19 disease, and healthcare acquired infection and AMR risk are still challenging for healthcare facilities that causing healthcare providers, clients, and cleaners are daily facing the risk of infection. The country needs to act now to ensure basic WASH services in at the healthcare setting. Hence, the study serves as an input for policymakers and programmers to design healthcare WASH frameworks and appropriate monitoring tools to tackle infection in healthcare settings. The finding also pledges leverage of adequate resources from partners, the ministry of health and healthcare facilities; and leadership commitment for the attainment of basic access to healthcare WASH services.

## Conclusions and Recommendation

The availability of healthcare WASH services in Addis Ababa city remains far short of the pace to achieve the SDG target (80% of facilities have basic services) by 2025. This study founds, based on the JMP service ladder none of the healthcare facilities in Addis Ababa city had basic access to all WASH services.

Limited access to WASH services and multiple existing challenges at healthcare facilities in Ethiopia makes worsening the prevention and control of COVID-19 pandemics, healthcare acquired infection, and AMR risk. It should be given priority and the need for more financial investment, capacity building, the commitment of leadership, and participation of partners in healthcare WASH service to ensure basic healthcare WASH and minimized the risk of health emergencies and AMR in healthcare settings.

## Data Availability

All relevant data are within the manuscript and its Supporting Information files

## Acknowledgements

We would like to acknowledge St. Paul Hospital Millennium Medical College Ethical Review Board and Addis Ababa City Administration Regional Health Bureau to assured ethical clearance letter. We also acknowledge all hospital and health centre management staff, for their commitment and information provided to conduct this study. Our thanks also extended to data collectors, supervisors, participants and friends contributed for the study.

